## Supplement 2 for "Racism in public health authorities – a scoping review and situational analysis"

**Supplementary material 2: Search protocols**

Searches conducted on 14.06.2022

Updated search conducted on 04.10.2024

### **Pubmed**

("infection protection"[All Fields] OR "STI"[All Fields] OR "STD"[All Fields] OR ("sexually transmitted diseases"[MeSH Terms] OR ("sexually"[All Fields] AND "transmitted"[All Fields] AND "diseases"[All Fields]) OR "sexually transmitted diseases"[All Fields] OR ("sexually"[All Fields] AND "transmitted"[All Fields] AND "disease"[All Fields]) OR "sexually transmitted disease"[All Fields]) OR ("hiv"[MeSH Terms] OR "hiv"[All Fields]) OR "tuberculosis"[All Fields] OR "screening"[All Fields] OR "sex work"[All Fields] OR "sex industry"[All Fields] OR "prostitution"[All Fields] OR "abortion"[All Fields] OR "pregnancy"[All Fields] OR "social psychiatry"[All Fields] OR "counselling"[All Fields] OR "counseling"[All Fields] OR "social work"[All Fields] OR "deportation"[All Fields]) AND ("racism"[MeSH Terms] OR "racism"[All Fields] OR "racist"[All Fields] OR "racial prejudice"[All Fields] OR "racial prejudices"[All Fields] OR "racial bias"[All Fields] OR "racial discrimination"[All Fields] OR "racial discrimination"[All Fields] OR "racialisation"[All Fields] OR "racialization"[All Fields] OR "racialised"[All Fields] OR "racialized"[All Fields] OR "racial microaggression"[All Fields] OR "racial stereotype"[All Fields] OR "racial abuse"[All Fields] OR "racial hostility"[All Fields] OR "racial harassment"[All Fields] OR "racial oppression"[All Fields] OR "racial equity"[All Fields] OR "anti-racism"[All Fields] OR "critical race theory"[All Fields]) AND ("public health"[All Fields] OR "health protection"[All Fields] OR "health security"[All Fields] OR "public health service"[All Fields] OR "public health administration"[All Fields] OR "public health authority"[All Fields] OR "public health authorities"[All Fields] OR "public health office"[All Fields] OR "public health department"[All Fields] OR "public health agency"[All Fields] OR "public health agencies"[All Fields] OR "public health institution"[All Fields])

**Translations**

**sexually transmitted disease:** "sexually transmitted diseases"[MeSH Terms] OR ("sexually"[All Fields] AND "transmitted"[All Fields] AND "diseases"[All Fields]) OR "sexually transmitted diseases"[All Fields] OR ("sexually"[All Fields] AND "transmitted"[All Fields] AND "disease"[All Fields]) OR "sexually transmitted disease"[All Fields]

**HIV:** "hiv"[MeSH Terms] OR "hiv"[All Fields]

**racism[MeSH Terms]:** "racism"[MeSH Terms]

### **EMBASE**

('infection protection' OR 'sti' OR 'std'/exp OR 'std' OR 'sexually transmitted disease'/exp OR 'sexually transmitted disease' OR (sexually AND transmitted AND ('disease'/exp OR disease)) OR 'hiv'/exp OR hiv OR 'tuberculosis'/exp OR 'tuberculosis' OR 'screening'/exp OR 'screening' OR 'sex work'/exp OR 'sex work' OR 'sex industry' OR 'prostitution'/exp OR 'prostitution' OR 'abortion'/exp OR 'abortion' OR 'pregnancy'/exp OR 'pregnancy' OR 'social psychiatry'/exp OR 'social psychiatry' OR 'counselling'/exp OR 'counselling' OR 'counseling'/exp OR 'counseling' OR 'social work'/exp OR 'social work' OR 'deportation'/exp OR 'deportation') AND ('racism'/exp OR 'racism' OR 'racist' OR 'racial prejudice'/exp OR 'racial prejudice' OR 'racial prejudices' OR 'racial bias'/exp OR 'racial bias' OR 'racial discrimination'/exp OR 'racial discrimination' OR 'racialisation' OR 'racialization' OR 'racialised' OR 'racialized' OR 'racial microaggression' OR 'racial stereotype' OR 'racial abuse' OR 'racial hostility'/exp OR 'racial hostility' OR 'racial harassment' OR 'racial oppression' OR 'racial equity' OR 'anti-racism' OR 'critical race theory') AND ('public health'/exp OR 'public health' OR 'health protection'/exp OR 'health protection' OR 'health security'/exp OR 'health security' OR 'public health service'/exp OR 'public health service' OR 'public health administration'/exp OR 'public health administration' OR 'public health authority' OR 'public health authorities' OR 'public health office' OR 'public health department' OR 'public health agency' OR 'public health agencies' OR 'public health institution') AND [embase]/lim

### **CINAHL**

( "infection protection" OR "STI" OR "STD" OR sexually transmitted disease OR HIV OR "tuberculosis" OR "screening" OR "sex work" OR "sex industry" OR "prostitution" OR "abortion" OR "pregnancy" OR "social psychiatry" OR "counselling" OR "counseling" OR "social work" OR "deportation" ) AND ( "racism" OR "racist" OR "racial prejudice" OR "racial prejudices" OR "racial bias" OR "racial discrimination" OR "racial discrimination" OR "racialisation" OR "racialization" OR "racialised" OR "racialized" OR "racial microaggression" OR "racial stereotype" OR "racial abuse" OR "racial hostility" OR "racial harassment" OR "racial oppression" OR "racial equity" OR "anti-racism" OR "critical race theory" ) AND ( "public health" OR "health protection" OR "health security" OR "public health service" OR "public health administration" OR "public health authority" OR "public health authorities" OR "public health office" OR "public health department" OR "public health agency" OR "public health agencies" OR "public health institution" )

**Expanders** - Apply related words; Also search within the full text of the articles; Apply equivalent subjects

**Search modes** - Boolean/Phrase

### **APA PSYCINFO**

( "infection protection" OR "STI" OR "STD" OR sexually transmitted disease OR HIV OR "tuberculosis" OR "screening" OR "sex work" OR "sex industry" OR "prostitution" OR "abortion" OR "pregnancy" OR "social psychiatry" OR "counselling" OR "counseling" OR "social work" OR "deportation" ) AND ( "racism" OR "racist" OR "racial prejudice" OR "racial prejudices" OR "racial bias" OR "racial discrimination" OR "racial discrimination" OR "racialisation" OR "racialization" OR "racialised" OR "racialized" OR "racial microaggression" OR "racial stereotype" OR "racial abuse" OR "racial hostility" OR "racial harassment" OR "racial oppression" OR "racial equity" OR "anti-racism" OR "critical race theory" ) AND ( "public health" OR "health protection" OR "health security" OR "public health service" OR "public health administration" OR "public health authority" OR "public health authorities" OR "public health office" OR "public health department" OR "public health agency" OR "public health agencies" OR "public health institution" )

**Expanders** - Apply related words; Also search within the full text of the articles; Apply equivalent subjects

**Search modes** - Boolean/Phrase
