## Supplement 3 for "Racism in public health authorities – a scoping review and situational analysis"

**Supplementary material 3: Data charting**

Table 3 Data charting items and supporting descriptions

| **Category** | **Description** |
| --- | --- |
| Reference | Full reference of the original article |
| Type of material | Study, commentary/editorial, dissertation, book chapter |
| Study design | Qualitative, quantitative or mixed-methods study, plus more detailed information on the study (e.g. cross-sectional study, focus groups) |
| Study population | Which study population is analysed in the material? |
| Geographic area of the study | Which country is analysed in the material? |
| Definition of racism | Has racism been defined in the material and if so, how? |
| Assessment of racism | How has racism been assessed? (e.g. specific tools, questionnaires? |
| Role of the public health department | What role of the public health department was explored? (e.g. counselling, preventive services incl. vaccination, surveillance & monitoring, policies development) |
| Involvement of racialised groups? | (How) did the material involve racialized groups when generating evidence? |
| Situational map | Assess what kind of elements and actants have been addressed, and how   - Individual human elements/ actants - Nonhuman elements/actants - Collective human elements/ actants - Implicated silent elements/actants - Discursive constructions of individual and/or collective actants - Discursive constructions of nonhuman actants - Political/economic elements - Sociocultural/symbolic elements - Temporal elements - Spatial elements - Major issues/debates (usually contested) - Related discourses (historical, narrative, and or visual) - Other kinds of elements |
